# Reinfection by the SARS-CoV-2 Gamma variant in blood donors in Manaus, Brazil

**DOI:** 10.1101/2021.05.10.21256644

**Authors:** Carlos A. Prete, Lewis F. Buss, Renata Buccheri, Claudia M. M. Abrahim, Tassila Salomon, Myuki A. E. Crispim, Marcio K. Oikawa, Eduard Grebe, Allyson G. da Costa, Nelson A. Fraiji, Maria do P. S. S. Carvalho, Charles Whittaker, Neal Alexander, Nuno R. Faria, Christopher Dye, Vítor H. Nascimento, Michael P. Busch, Ester C. Sabino

## Abstract

**Background:** The city of Manaus, north Brazil, was stricken by a second epidemic wave of SARS-CoV-2 despite high seroprevalence estimates, coinciding with the emergence of the Gamma (P.1) variant. Reinfections were postulated as a partial explanation for the second surge. However, accurate calculation of reinfection rates is difficult when stringent criteria as two time-separated RT-PCR tests and/or genome sequencing are required. To estimate the proportion of reinfections caused by the Gamma variant during the second wave in Manaus and the protection conferred by previous infection, we analyzed a cohort of repeat blood donors to identify anti-SARS-CoV-2 antibody boosting as a means to infer reinfection.

**Methods:** We tested serial blood samples from unvaccinated repeat blood donors in Manaus for the presence of anti-SARS-CoV-2 IgG antibody. Donors were required to have three or more donations and at least one donation during each epidemic wave. Donors were tested with two assays that display waning in early convalescence, enabling the detection of reinfection-induced boosting. The serial samples were used to divide donors into six groups defined based on the inferred sequence of infection and reinfection with non-Gamma and Gamma variants.

**Results:** From 3,655 repeat blood donors, 238 met all inclusion criteria, and 223 had enough residual sample volume to perform both serological assays. Using a strict serological definition of reinfection, we found 13.6% (95% CI 7.0% - 24.5%) of all presumed Gamma infections that were observed in 2021 were reinfections. If we also include cases of probable or possible reinfections, these percentages increase respectively to 22.7% (95% CI 14.3% - 34.2%) and 39.3% (95% CI 29.5% - 50.0%). Previous infection conferred a protection against reinfection of 85.3% (95% CI 71.3% - 92.7%), decreasing to respectively 72.5% (95% CI 54.7% - 83.6%) and 39.5% (95% CI 14.1% - 57.8%) if probable and possible reinfections are included.

**Conclusions:** Reinfection due to Gamma is common and may play a significant role in epidemics where Gamma is prevalent, highlighting the continued threat variants of concern pose even to settings previously hit by substantial epidemics.

## Introduction

Approximately 76% of the inhabitants of Manaus had been infected with SARS-CoV-2 eight months after the first reported case in March 2020[1]. Nevertheless, a second epidemic wave occurred in the city, coinciding with the emergence of a new SARS-CoV-2 variant of concern (VOC) in November 2020 first denoted P.1, and recently classified as the Gamma variant of concern by WHO[2]. This variant corresponded to 87% of all infections in January 2021[3].

Both increased transmissibility and the ability to partially evade protective immunity have been postulated to explain the Gamma-driven resurgence of COVID-19 in Manaus[3, 4]. Whilst a significant body of work supports increases to transmissibility of both Gamma and other variants of concern[5–7], comparatively little work has explored the potential for reinfection by these lineages, despite significant in vitro evidence supporting partial immune-escape[8]. It is therefore essential to understand the rate of reinfection in order to predict how this variant (and others with immune-escape potential) will spread through Brazil and other regions of the globe that have experienced significant previous outbreaks and are at risk for reinfections.

Individual cases of reinfection by the Gamma variant have been widely reported in the literature[9–11], but the frequency of these cases at a population-level has not been established. Detecting reinfections directly by testing of swabs from recurrent symptomatic infections is difficult because most SARS-CoV-2 infections are undiagnosed asymptomatic or mild cases[12], leading to a small number of patients with two confirmed infections. Instead, we assess the reinfection rate using samples of repeat blood donors, allowing the detection of asymptomatic infections.

## Methods

We retrieved and tested serial samples from unvaccinated repeat donors from Manaus with three or more donations, which included at least one during the first epidemic wave (between April 1^st^ and June 30^th^, 2020) and at least one after January 1^st^, 2021). We excluded donors that had their first anti-N positive result in November and December 2020, when it was not possible to determine whether the infection was caused by Gamma due to its low prevalence at that time. Given this exclusion, no infections observed in 2020 are due to Gamma because this VOC had an insignificant prevalence before November 2020[3]. The samples were first tested using an anti-N SARS-CoV-2 IgG chemiluminescence microparticle assay (CIMA, Abbott Park, IL, USA), and then the samples with enough volume were retested using the Abbott anti-S SARS-CoV-2 IgG CIMA. These are high specificity assays whose reactivity consistently wanes during convalescence[1, 13, 14] and presents small measurement error (see **Supplemental Appendix**). Furthermore, because the anti-S assay shows smaller reactivity waning than the anti-N assay, it may be able to detect infections that remained undetected by the anti-N assay.

In order to detect reinfections, we hypothesized that reinfection would induce anamnestic “boosting” of plasma anti-N and anti-S IgG antibody levels, yielding a V-shaped time series of antibody reactivity levels. We also assumed that a V-shaped antibody curve can only be caused by reinfection, since exposure should only lead to an increase of the antibody level if there is significant viral replication, hence an infection.

Repeat donors were partitioned into six groups that reflect the inferred sequence of infection and reinfection with non-Gamma and Gamma variant. To define these groups, we classified donors based on the serial anti-N samples, and we also obtained a second classification based on the serial anti-S samples. The groups assigned for each assay were then combined according to **Table 1** to obtain the final classifications. Donors that could not be classified because some of their samples did not have enough volume to be tested with the anti-S assay received the “Unknown” classification for the anti-S assay. Reinfections detected by only one assay were classified as possible reinfections, and only cases of reinfections detected by both assays had their final classification assigned as reinfection. Therefore, the rules for obtaining the final groups are conservative.

**Table 1.** Final classification for each donor based on the antibody pattern obtained with both the anti-N and anti-S assays, and the number of donors assigned to each group. Empty cells represent groups with no donors. Text within the cells denotes the final classification assigned to each case.

| Classification of donors<br>(anti-S assay) |  |  |  |  |  |  |  |
| --- | --- | --- | --- | --- | --- | --- | --- |
| Classification of donors<br>(anti-N assay) |  | Persistently<br>seronegative | Infection by<br>non-Gamma<br>variant | Infection<br>by Gamma | Reinfection by<br>Gamma | Probable<br>reinfection by<br>Gamma | Unknown |
|  | Persistently<br>seronegative | Persistently<br>seronegative<br>(60) | Infected by<br>non-Gamma<br>variant (4) | Infected by<br>Gamma (3) | Possible<br>reinfection by<br>Gamma (1) | .. | Persistently<br>seronegative (4) |
|  | Infection by<br>non-Gamma<br>variant | .. | Infected by<br>non-Gamma<br>variant (77) | .. | Possible<br>reinfection by<br>Gamma (14) | Infected by non-Gamma variant<br>(2) | Infected by non-Gamma variant<br>(7) |
|  | Infection by<br>Gamma | Infected by<br>Gamma (1) | .. | Infected by<br>Gamma<br>(43) | Possible<br>reinfection by<br>Gamma (1) | .. | Infected by<br>Gamma (4) |
|  | Reinfection<br>by Gamma | Possible<br>reinfection<br>by Gamma<br>(1) | .. | Possible<br>reinfection<br>by Gamma<br>(1) | Reinfection by<br>Gamma (8) | .. | .. |
|  | Probable<br>reinfection<br>by Gamma | .. | .. | .. | .. | Probable<br>reinfection by<br>Gamma (7) | .. |

To design the inclusion rules for each group, we assumed that all positive cases in 2021 are due to Gamma because of its high prevalence (87% of sequenced samples) in early January, which likely increased after January due to the higher transmissibility of Gamma compared to non-Gamma variants circulating in Manaus[3]. **Supplemental Figure 1** contains a flowchart illustrating the classification rules, and **Supplemental Figures 2 and 3** contain the serial results of each group for the anti-N and anti-S assays. The groups and their corresponding definitions are listed below, and are also summarized in **Supplemental Table 1**.

A. **Persistently seronegative** Donors that never had a positive donation. It is not possible to say that all persistently seronegative donors were not infected, since some infected donors may have had already seroreverted at the date of sample collection, or not seroconverted at all.
B. **Infection by non-Gamma variant** Two requirements are needed for a donor to be included in this group. First, the donor must have a positive donation before November 1^st^, 2020. Since donors that had their first positive result in November and December 2020 were excluded, this requirement is equivalent to requiring a positive donation in 2020 and a negative donation in 2021. Donors must also fill one of the following rules:
  a. All donations in 2021 are negative.
  b. There are positive donations in 2021, but none of them have a rising result.
C. **Infection by Gamma** Donors that did not have any positive donation in 2020, but had a positive donation in 2021. Some of these cases may be unobserved reinfections by Gamma in the case of an undetected infection in 2020.
D. **Reinfection by Gamma** A donor is classified as a case of reinfection by Gamma if any of the following rules if fulfilled:
  a. Donors with a positive donation in 2020 and another positive donation in 2021 with a V-shaped curve ending in 2021. In other words, these are donors that have a positive donation in 2020, a second donation with lower S/C value (that could be positive or negative), followed by a positive donation in 2021 with an increase in S/C value. These donors were seroreverting and then seroconverted again due to the reinfection.
  b. Donors with three consecutive rising positive results, the last being in 2021. Since the minimum interval between successive donations is 60 days for men and 90 days for women in Brazil, donors with three consecutive rising positive results would apparently be seroconverting for more than 120 days > Δ*t*_min_, a possibility that we rule out due to the definition of Δ*t*_min_. Donors following this rule have likely had an unobserved S/C decay after the second rising result, but seroconverted again after being reinfected.
E. **Probable reinfection by Gamma** Donors with two consecutive rising positive results, the last being in 2021, separated by an interval Δ*t* ≥ Δ*t*_min_, where Δ*t*_min_ is a predefined parameter equal to 141 days and 126 days for the anti-N and anti-S assays respectively (see **Supplemental Appendix** for an explanation on how Δ*t*_min_ was obtained). We hypothesize that donors following this rule have had an unobserved antibody decline after the first positive sample, and then seroconverted again after being reinfected. A minimum interval between donations is required to avoid misclassifying donors sampled during the seroconversion period as probable reinfections.

These rules imply that a truly reinfected individual may be misclassified as “Infection by Gamma” or “Infection by non-Gamma variant” if samples are not collected shortly after infection of reinfection, underestimating the proportion of reinfections. This effect is illustrated in **Figure 2**, which shows the idealized signal-to-cutoff curve of a reinfected individual and the corresponding classification based on the sequence of dates of sample collection.

**Figure 1.**
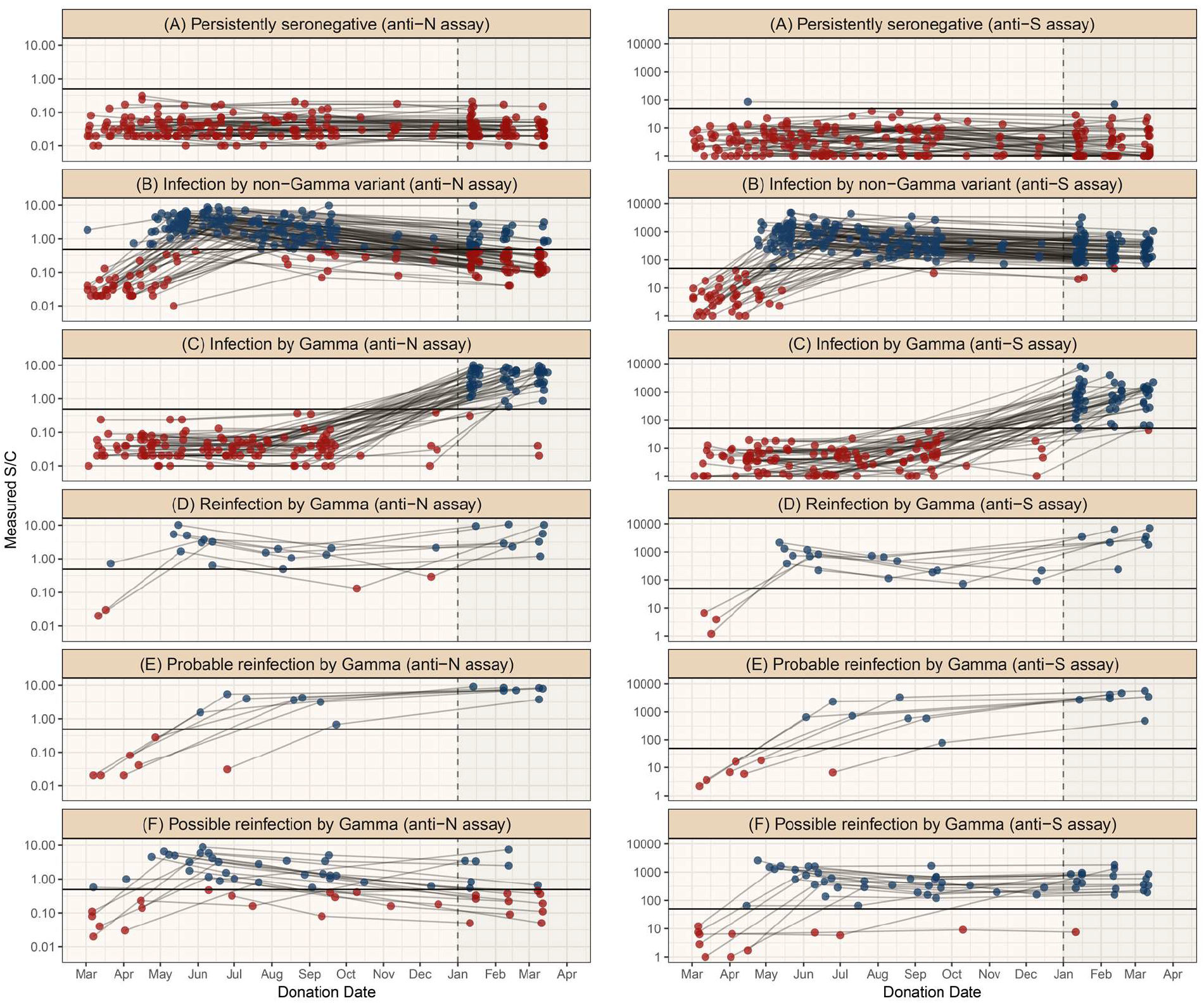
Classification of the repeat blood donors according to their antibody profile. Each facet shows the serial results obtained with the anti-N or anti-S IgG assays for donors in the corresponding group. Blue and red dots represent respectively positive and negative results, and donations from the same donor are connected by a line. Because 18 samples could not be retested with the anti-S assay, less than three anti-S results are shown for some donors, which were classified based solely on the serial anti-N results.

**Figure 2.**
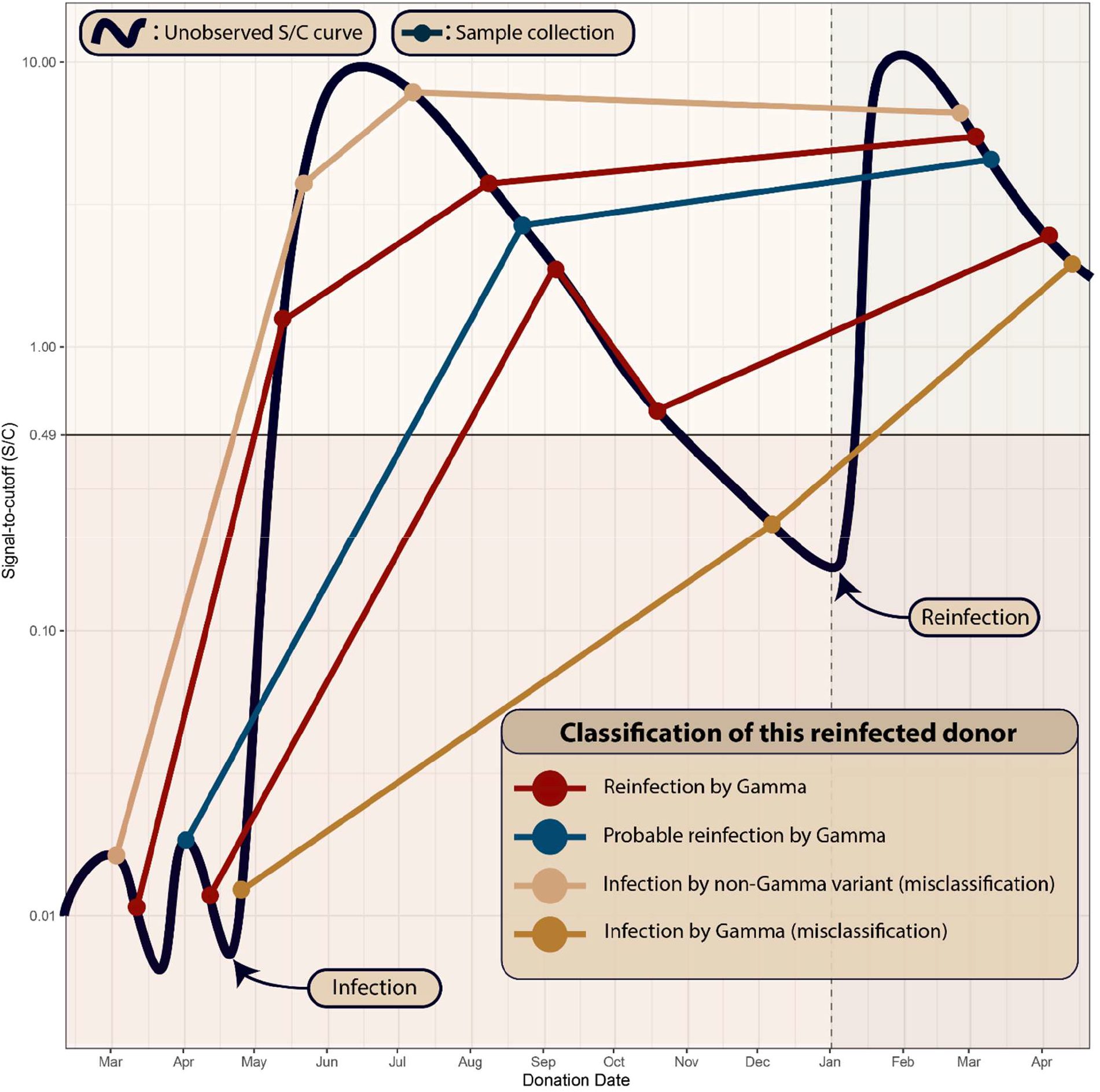
Illustration of an idealized signal-to-cutoff (S/C) curve of a reinfected individual that is assigned to different groups depending on the sequence of dates of sample collection. The black curve represents the unobserved trajectory of S/C over time, and circles represent sample collections. This figure shows five sets of serial samples that were collected in different dates. The patterns that can be confidently attributed to reinfection are shown in red: sampled points that reveal the underlying V-shaped curve, or three consecutive rising values that can only be obtained by sampling the underlying V-shaped curve. If the dates of sample collection are too sparse, this reinfection individual may be misclassified as “Infection by non-Gamma variant” or “Infection by Gamma”.

## Results

During the study period, we identified 3,655 repeat blood donors, of which 240 met our inclusion criteria (see **Supplemental Figure 1**). Two donors were excluded for having their first anti-N positive result in November or December 2020, resulting in 238 donors selected for this study. The median (IQR) age was 36.5 (28.0 – 44.0) and 12.8% were female. 18 samples were tested only with the anti-N assay because they did not have enough volume to be tested by the second assay. For this reason, only 223 donors were classified based on the anti-S assay patterns over time, and the remaining 15 donors were classified based solely on the anti-N assay. **Table 1** presents the results obtained by both assays, which showed a high concordance, with 87.4% of donors receiving the same classification on both assays. **Table 2** summarises the definition of groups and contains the sizes of the final groups, as well as the sizes of the groups for each assay. There were 59 presumed Gamma infections in 2021, of which 8 (13.6%, 95% CI 7.0% - 24.5%) had a V-shaped curved indicating reinfection by both anti-N and anti-S assays. The anti-S assay detected 16 cases of reinfection that were undetected by the anti-N assay; these were given a final classification of possible reinfection. If probable and possible reinfections are included, these percentages increase to 22.7% (95% CI 14.3% - 34.2%), or 39.3% (95% CI 29.5% - 50.0%), respectively. These 8 Gamma reinfections also represent 6.5% (95% CI 3.3% - 12.3%) of the 123 individuals that had a primary infection in the first wave, increasing to 12.2% (95% CI 7.5% - 19.1%) and 26.8% (95% CI 19.8% - 35.3%) if probable and possible reinfections are considered.

**Table 2.** Summarised definition and size of the groups used to classify donors for each assay. The final classification was obtained by combining the groups assigned by both assays according to **Table 1**. The definitions of probable reinfections depend on the parameter Δ*t*_min_ = 141 days for the anti-N assay and 126 days for the anti-S assay.

| Infection Group | Definition | Number of donors |  |  |
| --- | --- | --- | --- | --- |
|  |  | Anti-N assay | Anti-S assay | Final Classification |
| Persistently seronegative | No positive results | 72 | 62 | 64 |
| Infection by non-Gamma variant | A positive result before Nov 1<sup>st</sup> 2020 and decaying antibody levels in 2021. | 100 | 81 | 90 |
| Infection by Gamma | No positive results in 2020 and a positive result in 2021 | 49 | 47 | 51 |
| Reinfection by Gamma | A positive result in 2021 and before Nov 1<sup>st</sup> 2020 and an intermediate result with value below these two readings (V-shaped S/C time series). | 9 | 22 | 8 |
|  | A positive donation in 2021 succeeding two consecutive rising positive results in 2020. Donors in this group have an observed seroconversion period much longer than expected. | 1 | 2 | 0 |
| Probable reinfection by Gamma | One positive result in 2020 and a higher positive result in 2021 separated by an interval of at least $\Delta t_{\min}$ (seroconversion period longer than expected). | 7 | 9 | 7 |
| Possible reinfection by Gamma | Classification as “Reinfection by Gamma” for only one assay. | .. | .. | 18 |
| Unknown (anti-S assay only) | Not enough volume to retest the sample with the anti-S assay. | .. | 15 | .. |
| **Total** | .. | 238 | 238 | 238 |

Of 115 previously negative individuals, 51 (44.3%, 95% CI 35.6% - 53.5%) were infected by Gamma over the time-period considered. As such, the protection against reinfection conferred by previous infection (defined as 100 × [1 – relative risk of reinfection]) is 85.3% (95% CI 71.3% - 92.7%), or 72.5% (95% CI 54.7% - 83.6%) and 39.5% (95% CI 14.1% - 57.8%) if probable or possible reinfections are included.

## Discussion

The high proportion of Gamma reinfections suggests that reinfection with this variant is common and may play a significant role in regions where Gamma is highly prevalent. The estimated relative risk of reinfection shows that even though a previous infection decreases the chance of reinfection, the protection is not close to 100%. Hence, immunity against non-Gamma variants achieved by natural infection may not prevent new outbreaks caused by Gamma. This conclusion may also extend to other variants where *in vitro* results support immune-escape potential equal or larger than Gamma.

Our proportion of reinfections by Gamma is compatible with a previous estimate of 28% obtained from a compartmental model[15] in which Manaus had an estimated prevalence of 78% in November 2020. On the other hand, the obtained relative risk of reinfection is higher than reported in the literature for non-Gamma variants[16–18], especially if we assume that part of the donors classified as probable and possible reinfections were reinfected. The protection conferred by previous infection estimated from a cohort of healthcare workers in United Kingdom[17] that were submitted to regular PCR tests was 84%, similar to the protection of 80.5% obtained from a cohort of confirmed COVID-19 cases in Denmark[16]. In a cohort of confirmed cases in Italy[18], only 0.31% of the previously positive individuals were reinfected, compared to 3.9% of primary infections for the negative cohort. However, confirmed cases are biased towards symptomatic individuals, which are likely to have a smaller reinfection risk than asymptomatic or oligosymptomatic individuals. Also, because the sensitivity of serological assays depends on the disease severity [19], traditional methods may not detect reinfections following a mild infection.

The main limitation of this study is that donors were not sampled frequently enough to robustly detect cases of reinfection, leading to the possible existence of undetected cases of reinfection. We attempted to resolve this issue by classifying the degree of evidence and identifying probable and possible reinfections. Further, repeat negative donors may not represent truly unexposed individuals, since not all PCR+ individuals produce antibodies to nucleocapsid proteins[20] and because sparse sampling may have resulted in missing the positive interval. Another limitation is that a small proportion of infections in 2021 were not caused by Gamma, thus our results may be slightly affected by the reinfection rate of the non-Gamma variants. Also, blood donors are biased towards asymptomatic and mild infections; therefore, our reinfection rates cannot be extrapolated to persons with more severe primary disease. Finally, we could not determine the clinical relevance of COVID-19 reinfection because we did not have access to previous signs and symptom information.

## Conclusions

Our data suggest that reinfection due to Gamma is common and more frequent than has been detected by traditional approaches. The estimated reinfection rates suggest that the Gamma variant may induce a higher reinfection risk than previous non-Gamma variants. Overall, our results reinforce concerns over the risk of reinfection particularly as variants continue to evolve, and demonstrate that repeat blood donor serosurveillance is valuable for documenting rates and correlates of reinfection that complement surveillance for reinfections or vaccine breakthrough infections based on serial swab-based surveillance programs.

## Data Availability

Anonymized individual-level data of the 238 selected blood donors along with a data dictionary is available at https://github.com/carlosprete/reinfection_manaus. Data related to all repeat blood donors can be shared upon request.

## Supplementary Appendix

### Materials and Methods

#### 1. Definition of the groups of donors

From all 3,655 repeat blood donors tested with the anti-N assay, we selected only donors with three or more donations because it is not possible to infer reinfection based on two time points. We also required donors to have one donation between March 1st, 2020 and June 30th, 2020, and one donation after January 1st, 2021. This is because most infections in the first wave happened between March and June, thus this requirement helps avoiding selecting donors that had their first sample collected many months after the date of infection, which may have a false negative result if they have already seroreverted when their first sample was collected. If this requirement is not employed, some cases of reinfection may be misclassified as infection by Gamma because the first infection was not detected. It is worth noting that this requirement depends only on the date of donation, and does not add a bias towards positive or negative results.

240 donors met these criteria and were tested with the anti-N assay. We excluded two donors who had their first positive anti-N result in November or December 2020 (when the prevalence of Gamma was small, but rising) because it is not possible to determine if they were infected by Gamma or a non-Gamma variant. The samples from the 238 selected donors were retested with the anti-S assay, except for 18 samples that did not have enough volume to be retested, causing 15 donors to be classified as “Unknown” for the anti-S assay.

The 238 and 223 selected donors for the anti-N and anti-S assays respectively were divided into five groups for each assay, and these groups were then combined according to **Table 1** to obtain the final classification for each donor. The definition of these groups depends on a predefined parameter Δ*t*_min_ used to define the expected behavior of non-reinfected individuals. This parameter represents the minimum interval between donations necessary to accept a probable reinfection, and it is estimated based on donations that occurred before the incidence of Gamma became significant (i.e., donations up to and including October 2020).

The objective of defining Δ*t*_min_ is to avoid misclassifying donors as reinfected when samples were collected during the seroconversion period – that is, we consider that Δ*t*_min_ is much greater than the period of seroconversion. Before estimating these parameters, we added to all donors an artificial negative donation with CIMA result 0.01 S/C on February 28, 2020, before the beginning of the epidemic in Manaus. This is because at that date SARS-CoV-2 had not yet been introduced to the population, which was presumably completely immunologically naïve at that time.

Let *N*(Δ*t*_i_) be the number of donors that have at least one pair of successive positive results before November 2020 separated by an interval Δ*t* ≥ Δ*t*_*i*_. The function *N*(Δ*t*_*i*_) represents the number of possible reinfections observed in 2020, for a given choice of Δ*t*_*i*_. We first estimate Δ*t*_min_ as the smallest interval Δ*t*_*i*_ such that *N*(Δ*t*_*i*_) = 0, obtaining Δ*t*_min_ = 141 days and 126 days for the anti-N and anti-S assays respectively. It is worth noting that changing the value of Δ*t*_min_ does not substantially change the number of probable reinfections because all cases of probable reinfections have samples separated by a large interval.

#### 2. Assessing the measurement error of the SARS-Cov-2 anti-N IgG chemiluminescence microparticle assay

We define the CIMA test to be positive if the measured signal-to-cutoff (S/C) is higher or equal to 0.49 for the anti-N assay. This is the lowest value of range defined by the manufacturer (CIMA, Abbott Park, IL, USA) and provides a specificity of 97.6% (95% CI 96.3% - 98.5%) based on 20 false-positives in 821 pre-pandemic blood donation samples in Manaus, and a peak sensitivity (prior to waning) of 91.7% (95% CI 87.0 – 94.4) based on 177 positive samples out of 193 PCR-positive symptomatic convalescent plasma donors tested 20-50 days following symptom onset[1]. For the anti-S assay, we use the cutoff of 50.0 recommended by the manufacturer to determine if the test is positive. In our analyses, we do not apply any correction based on the sensitivity and specificity of the assay.

Even though the anti-N assay has high sensitivity and specificity, it produces results that are subject to measurement error, which results in variation in S/C that does not reflect a biological change, but is simply variation within the limit of precision of the test. If this variation is not small, sequential donations may have a V-shaped curve even if reinfection has not occurred, leading to an overestimation of the reinfection rate. To assess the amount of measurement error, we tested 200 samples in replicate from blood donors that donated in February 2021 in São Paulo.

Supplemental Figure 4 shows the measured S/C for the first and the second test of each sample. The absolute deviation of each pair of measured S/C had a median of 0.00 and a 95% confidence interval of 0.00 - 0.09. If only positive results were considered, the median deviation increases to 0.02 (95% CI 0.00 - 0.16), and the relative deviation obtained by dividing the absolute deviation by the first result has median 1.21% (95% CI 0.00% - 7.3%) for positive results.

Therefore, the assay employed in this study yields results with a small amount of measurement error. For this reason, a sequence of serial samples is unlikely to be misclassified as a case of reinfection due to measurement noise.

The measurement error was not assessed for the anti-S assay, but this does not affect the robustness of our results because we used the anti-S assay as a secondary validation of the results obtained with the anti-N assay. Nevertheless, the data presented by Germanio et al[2] shows that the S/C measured with the anti-S assay consistently wanes over time, suggesting a small noise level as well.

**Supplemental Figure 1.**
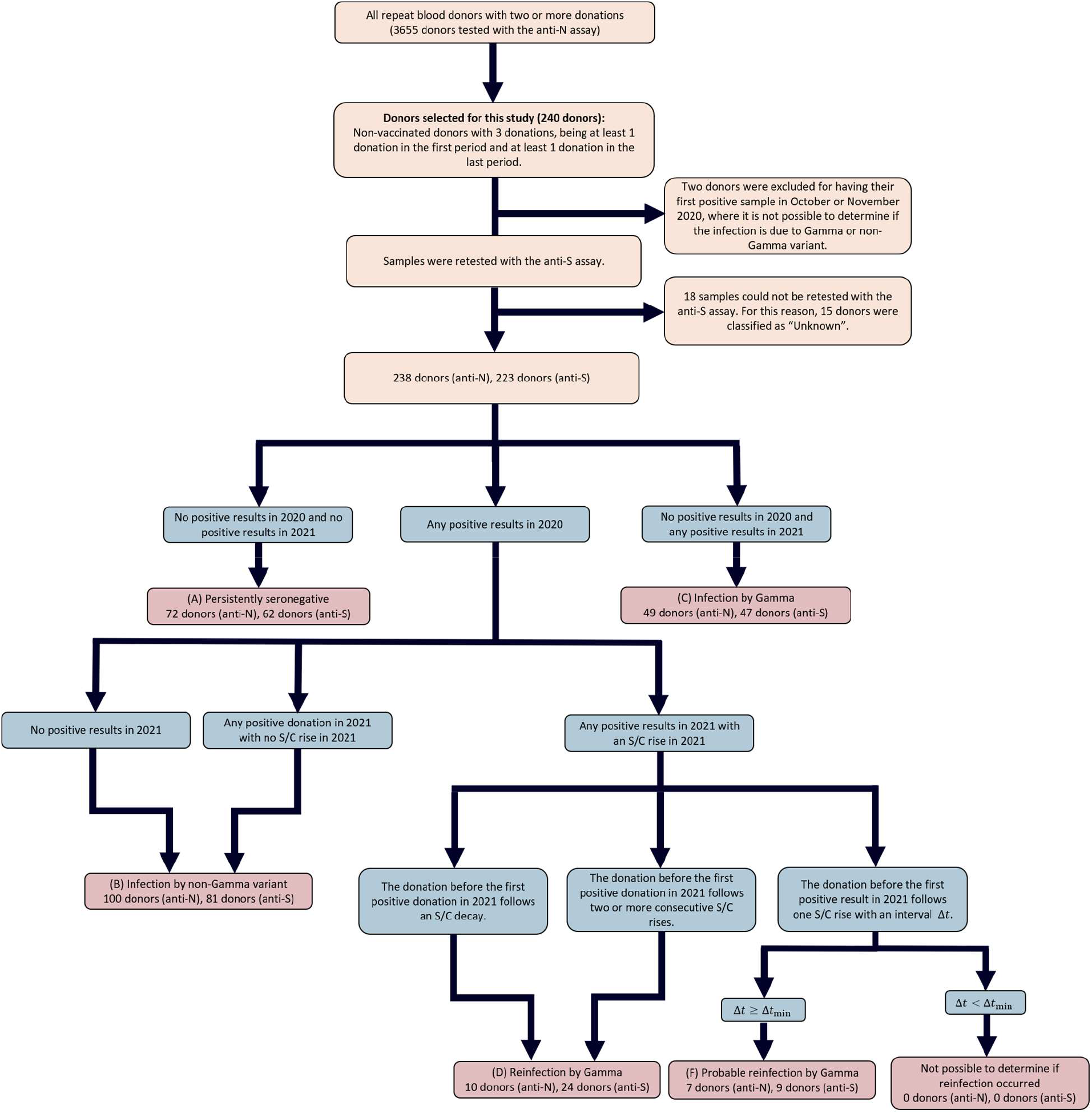
Flowchart describing how repeat blood donors were classified into the groups shown in Figure 1. We used Δ*t*_min_ = 141 days and Δ*t*_min_ = 126 days for the anti-N and anti-S assays respectively.

**Supplemental Figure 2.**
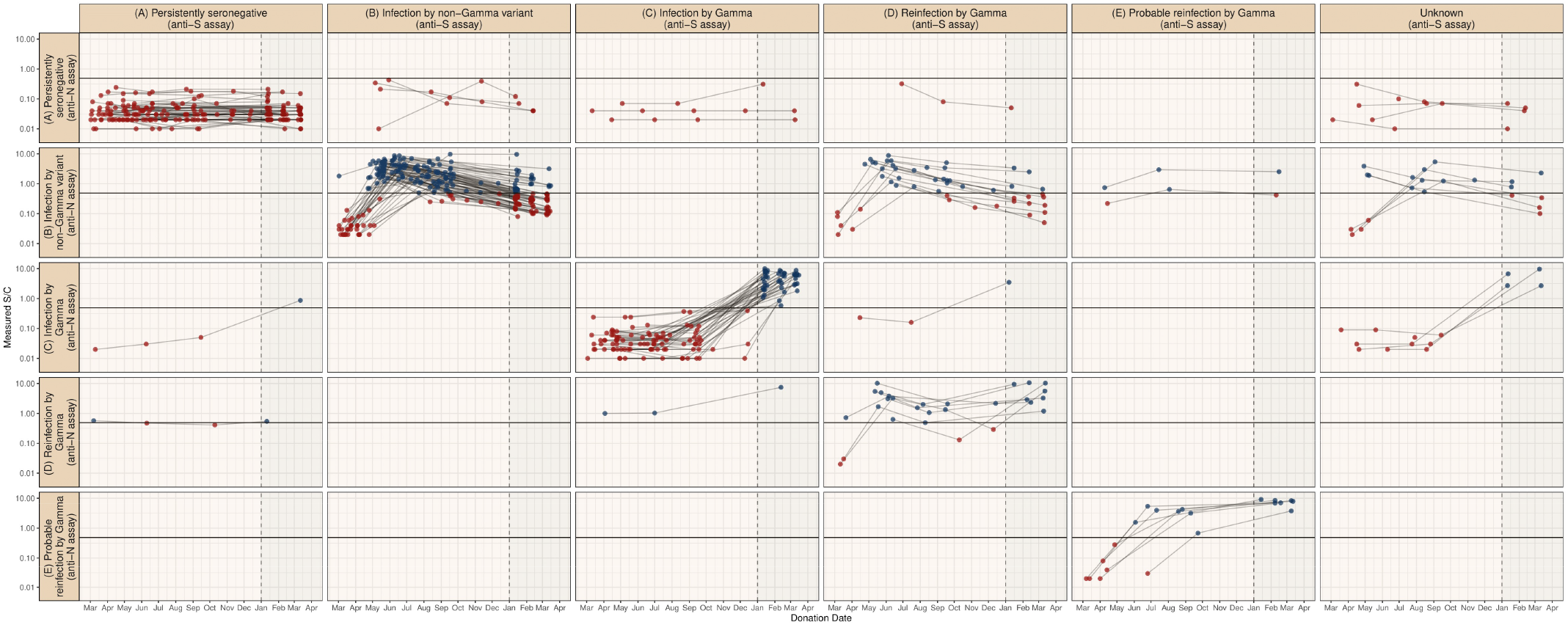
Serial results obtained with the anti-N assay for each group of donors.

**Supplemental Figure 3.**
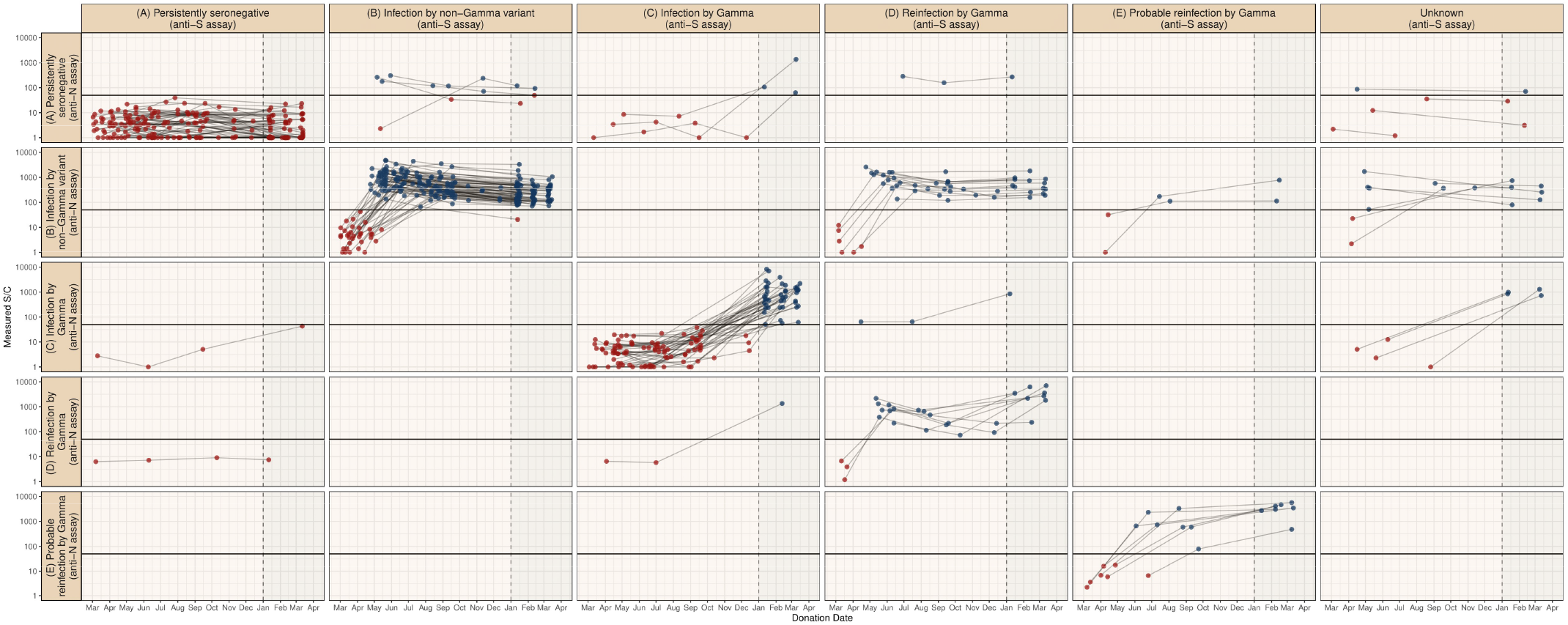
Serial results obtained with the anti-S assay for each group of donors.

**Supplemental Figure 4.**
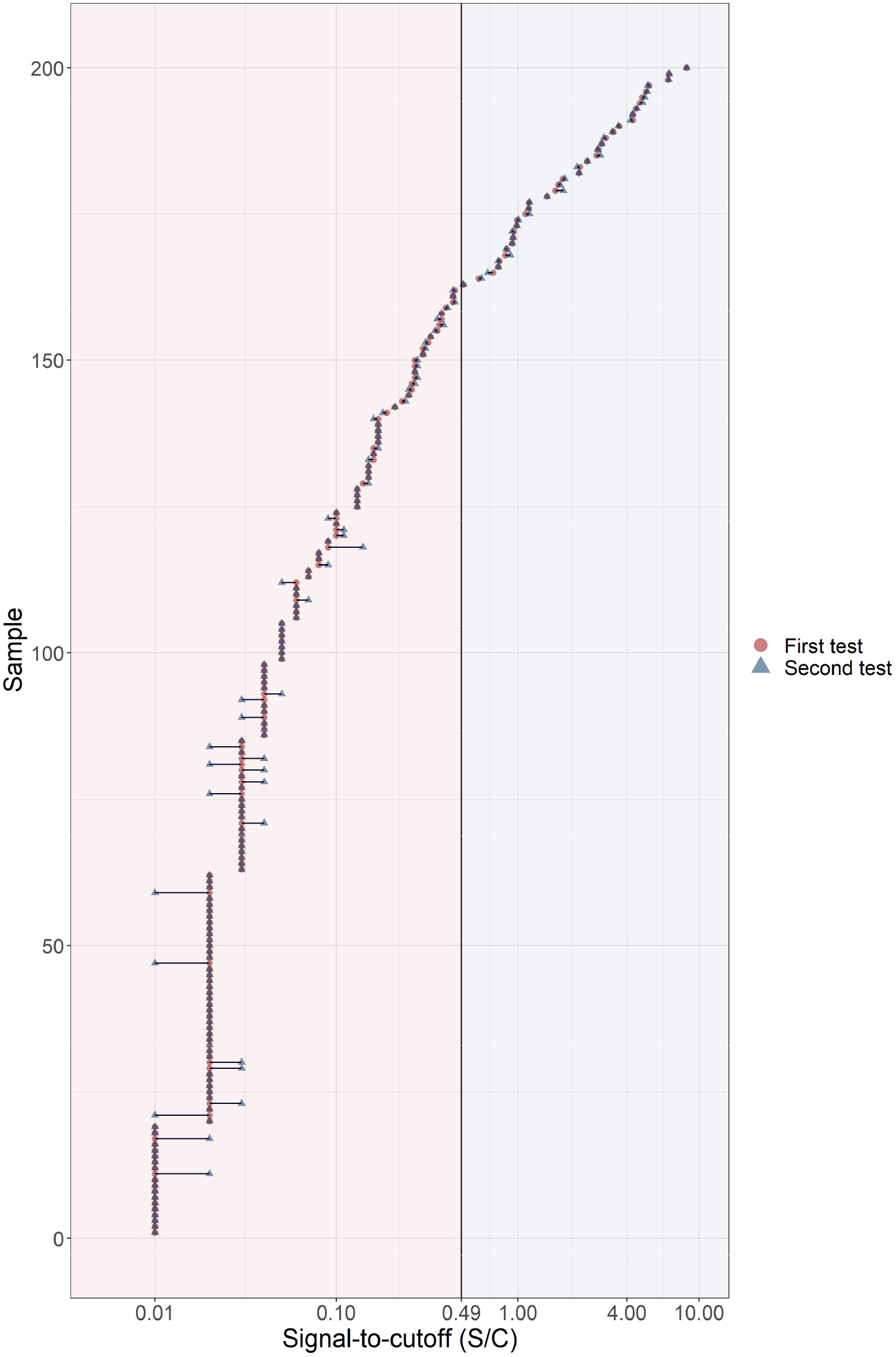
Validation of the noise level of the SARS-Cov-2 anti-N IgG chemiluminescence microparticle assay by testing 200 samples in replicate. Results corresponding to the same sample are connected by a horizontal line. The assay produces consistent results with very little variation.

## References

1. Buss LF, Prete CA, Abrahim CMM, Mendrone A, Salomon T, De Almeida-Neto C, et al. Three-quarters attack rate of SARS-CoV-2 in the Brazilian Amazon during a largely unmitigated epidemic. Science (80-). 2021;371:288–92. doi:10.1126/science.abe9728.

2. Tracking SARS-CoV-2 variants. https://www.who.int/en/activities/tracking-SARS-CoV-2-variants/. Accessed 31 Jul 2021.

3. Faria NR, Mellan TA, Whittaker C, Claro IM, Candido D da S, Mishra S, et al. Genomics and epidemiology of the P.1 SARS-CoV-2 lineage in Manaus, Brazil. Science (80-). 2021;:eabh2644. doi:10.1126/science.abh2644.

4. Sabino EC, Buss LF, Carvalho MPS, Prete CA, Crispim MAE, Fraiji NA, et al. Resurgence of COVID-19 in Manaus, Brazil, despite high seroprevalence. The Lancet. 2021;397:452–5. doi:10.1016/S0140-6736(21)00183-5.

5. Campbell F, Archer B, Laurenson-Schafer H, Jinnai Y, Konings F, Batra N, et al. Increased transmissibility and global spread of SARS-CoV-2 variants of concern as at June 2021. Eurosurveillance. 2021;26:2100509. doi:10.2807/1560-7917.ES.2021.26.24.2100509.

6. Volz E, Mishra S, Chand M, Barrett JC, Johnson R, Geidelberg L, et al. Assessing transmissibility of SARS-CoV-2 lineage B.1.1.7 in England. Nat 2021 5937858. 2021;593:266–9. doi:10.1038/s41586-021-03470-x.

7. Brown KA, Tibebu S, Daneman N, Schwartz K, Whelan M, Buchan S. Comparative Household Secondary Attack Rates associated with B.1.1.7, B.1.351, and P.1 SARS-CoV-2 Variants. medRxiv. 2021;:2021.06.03.21258302. doi:10.1101/2021.06.03.21258302.

8. Lucas C, Vogels CBF, Yildirim I, Rothman JE, Lu P, Monteiro V, et al. Impact of circulating SARS-CoV-2 variants on mRNA vaccine-induced immunity in uninfected and previously infected individuals. medRxiv. 2021;:2021.07.14.21260307. doi:10.1101/2021.07.14.21260307.

9. Naveca F, Costa C da, Nascimento V, al. et. SARS-CoV-2 reinfection by the new variant of concern (VOC) P.1 in Amazonas, Brazil. Virological. 2021.

10. Adrielle dos Santos L, Filho PG de G, Silva AMF, Santos JVG, Santos DS, Aquino MM, et al. Recurrent COVID-19 including evidence of reinfection and enhanced severity in thirty Brazilian healthcare workers. J Infect. 2021;82:399–406. doi:10.1016/j.jinf.2021.01.020.

11. Romano CM, Felix AC, Paula AV de, Jesus JG de, Andrade PS, Cândido D, et al. SARS-CoV-2 reinfection caused by the P.1 lineage in Araraquara city, Sao Paulo State, Brazil. Rev Inst Med Trop Sao Paulo. 2021;63. doi:10.1590/S1678-9946202163036.

12. Silva AAM da, Lima-Neto LG, Azevedo C de Mpes de, Costa LMM da, Bragança MLBM, Barros Filho AKD, et al. Population-based seroprevalence of SARS-CoV-2 and the herd immunity threshold in Maranhão. Rev Saude Publica. 2020;54:131. doi:10.11606/s1518-8787.2020054003278.

13. Lumley SF, Wei J, O’Donnell D, Stoesser NE, Matthews PC, Howarth A, et al. The duration, dynamics and determinants of SARS-CoV-2 antibody responses in individual healthcare workers. Clin Infect Dis. 2021. doi:10.1093/cid/ciab004.

14. Germanio C Di, Simmons G, Kelly K, Martinelli R, Darst O, Azimpouran M, et al. SARS-CoV-2 antibody persistence in COVID-19 convalescent plasma donors: Dependency on assay format and applicability to serosurveillance. Transfusion. 2021. doi:10.1111/TRF.16555.

15. Mendes Coutinho R, Maria F, Marquitti D, Ferreira LS, Borges ME, Lopes R, et al. Model-based estimation of transmissibility and reinfection of SARS-CoV-2 P.1 variant. medRxiv. 2021;:2021.03.03.21252706. doi:10.1101/2021.03.03.21252706.

16. Hansen CH, Michlmayr D, Gubbels SM, Mølbak K, Ethelberg S. Assessment of protection against reinfection with SARS-CoV-2 among 4 million PCR-tested individuals in Denmark in 2020: a population-level observational study. Lancet. 2021;397:1204–12. doi:10.1016/S0140-6736(21)00575-4.

17. Hall VJ, Foulkes S, Charlett A, Atti A, Monk EJ, Simmons R, et al. SARS-CoV-2 infection rates of antibody-positive compared with antibody-negative health-care workers in England: a large, multicentre, prospective cohort study (SIREN). Lancet. 2021;397:1459–69. doi:10.1016/s0140-6736(21)00675-9.

18. Vitale J, Mumoli N, Clerici P, De Paschale M, Evangelista I, Cei M, et al. Assessment of SARS-CoV-2 Reinfection 1 Year After Primary Infection in a Population in Lombardy, Italy. JAMA Intern Med. 2021. doi:10.1001/jamainternmed.2021.2959.

19. Peluso MJ, Takahashi S, Hakim J, Kelly JD, Torres L, Iyer NS, et al. SARS-CoV-2 antibody magnitude and detectability are driven by disease severity, timing, and assay. medRxiv Prepr Serv Heal Sci. 2021;:2021.03.03.21251639. doi:10.1101/2021.03.03.21251639.

20. Petersen LR, Sami S, Vuong N, Pathela P, Weiss D, Morgenthau BM, et al. Lack of Antibodies to Severe Acute Respiratory Syndrome Coronavirus 2 (SARS-CoV-2) in a Large Cohort of Previously Infected Persons. Clin Infect Dis. 2020. doi:10.1093/cid/ciaa1685.

## References

2. Germanio C Di, Simmons G, Kelly K, Martinelli R, Darst O, Azimpouran M, et al. SARS-CoV-2 antibody persistence in COVID-19 convalescent plasma donors: Dependency on assay format and applicability to serosurveillance. Transfusion. 2021. doi:10.1111/TRF.16555.

